# Telemedicine adoption in Ecuador: an assessment of physician perceptions and knowledge towards its benefits and limitation

**DOI:** 10.1101/2023.07.14.23292687

**Authors:** Ivan Cherrez-Ojeda, Emanuel Vanegas, Miguel Felix, María José Farfán Bajaña, Geovanny Alvarado-Villa, Hans Mautong, Fernando Espinoza, Zouina Sarfraz, Azza Sarfraz, Karla Robles-Velasco, Jack Michel, Abdelilah Lahmar, Luc J. I. Zimmermann, Antonio WD Gavilanes

## Abstract

Telemedicine is a growing field with the potential to improve healthcare delivery, however it is important for a proper implementation to understand how physicians perceive the benefits and limitations related to their use. With this study our aim is to assess the perceptions and knowledge of telemedicine among a sample of Ecuadorian healthcare providers. We conducted a cross-sectional online survey-based study where participants answered a 12-item survey assessing their knowledge and perceptions towards telemedicine. Demographic variables were analyzed applying descriptive statistics, and a chi-square goodness of fit test was used to assess the observed frequencies of each of the survey’s queries. In total, 382 participants completed the survey with an average age of 51.3 years (SD, 11.4). Around half of participants expressed to be lowly to very lowly familiarized with telemedicine technology (χ2(4) = 88.497, p = .000). Most of them considered to a high extent that telemedicine is effective in reducing costs of patient care in hospitals (32.5%; n=124; χ2(4) = 78.812, p = .000). Finally, 8 out 10 participants expressed that a framework should be created to prevent breaches of data confidentiality when using telemedicine (χ2(4) = 250.749, p = .000). In this study we found a considerable proportion of physicians reporting low familiarization with telemedicine despite being aware of the benefits it can bring to patient care. Breaches of data confidentially and the potential for malpractice were cited as main concerns in need of a framework to prevent them. Future studies are needed to address the perceived barriers of technology to ensure a safe and efficient use of telemedicine in the healthcare setting.

## Introduction

Telemedicine (TM) is defined as the remote clinical care of patients utilizing telecommunication technology as a substitute to face-to-face contact.(1) Its implementation can be seen across the medical field for the management of chronic lung disease, diabetes, high blood pressure, neurological disorders, dermatological conditions, oncology, and mental health problems over time.(2–5) Physicians can also benefit from telemedicine models by strengthening connections between health-care professionals to improve ongoing medical education and reducing professional isolation.(2)

Telemedicine is expected to bring various benefits, especially to those countries where the majority of the population live in remote areas without access to basic healthcare.(6,7) It is a growing field which has the potential to improve patient care, but also has many challenges associated with its adoption and usage. For instance, some studies have found that the implementation of technology may be negatively impacted by user perceived barriers.(8,9) Furthermore, a previous systematic review identified the expenses of technology and lack of literacy as major obstacles interrupting the successful implementation of telemedicine. (8) Therefore, identifying the main barriers among patients and healthcare providers is a key step for successful implementation of technology that needs to be addressed with additional studies.

Notably, the perceptions and knowledge concerning telemedicine vary among healthcare providers, with beliefs differing between users and non-users.(10) Factors including lack of technical expertise, and insufficient integration of data for continuity of care are notable barriers between clinicians and telemedicine.(11) As previously described it can lead to a situation in which despite widespread use of availability of telemedicine, the limited knowledge of clinicians regarding these technologies could potentially lead to underutilization of the resources.(12) Consequently, we have designed this study to assess the perceptions and knowledge of telemedicine among a sample of Ecuadorian healthcare providers (HCPs).

## Methods

### Study design and participants

We conducted a cross-sectional online survey-based study using a non-probability, snowball-sampling method where recruited participants provided referrals to recruit other HCPs. The sample comprised HCPs working in Ecuador with an active medical practice. The participants anonymously responded the 12-item survey using a Likert-scale.

### Questionnaire

For this cross-sectional study, the questionnaire by Ayatollahi *et al*. was previously adapted and validated for its use in the Ecuadorian HCPs population. (12,13) The survey consisted of two parts. The first part included the demographic information of participants (age, gender, higher education degree, medical specialty, and work experience). The second part of the survey included 4 domains where participants described their knowledge and perceptions about TM as follows:

1. Knowledge about Telemedicine (KAT): Q_1_-Q_3_
2. Perception of the Utility of Telemedicine (PUT): Q_4_-Q_6_
3. Perception of the Disadvantages of Telemedicine (PDT): Q_7_-Q_9_
4. Knowledge of the Security of Telemedicine (KST): Q_10_-Q_12_.

Each domain had 3 questions, for a total of 12 questions. Each question was answered based on a five-point Likert scale that ranged from very low (1) to very high (5). Each of the questions in the survey according to their domains can be visualized in the **S1 Table**.

### Statistical analysis

The present study reports demographic characteristics applying descriptive statistics. Nominal variables are presented as frequencies and percentages, while normally distributed continuous data is summarized through means and standard deviations. For the purposes of analysis, the 5-point Likert scale was used as a categorical variable, as a chi-square goodness of fit test was used to assess if the observed frequencies of each of the survey’s queries responses were as expected by chance or not. In case of assumption violation, a Fisher’s exact test was applied. All statistical analyses were performed using SPSS for Windows (version 25.0; SPSS Inc, Chicago, Illinois). Statistical significance was considered as *p*<0.05.

### Ethical considerations

This study was approved by “ Comité de ética e Investigación en Seres Humanos” (CEISH), ethical review board, Kennedy Hospital, Guayaquil-Ecuador (#HCK-CEISH-18-0060). Additionally, as per the Declaration of Helsinki, written informed consent was obtained from all participants.

## Results

### Descriptive statistics of demographics

In total, 382 participants completed the survey, with a response rate of 95%. The average age of the sample was 51.3 years (SD, 11.4); 58.6% were males. While 82.5% (n=315) of the participants had a specialty, only one in ten (10.2%; n=39) had a higher education degree. Regarding specialties, pediatrics (28.0%; n=107) and traumatology (13.1%, n= 50) were the most common. More than half (53.4%, n=204) of the participants had more than 20 years of work experience. Demographic characteristics are best summarized in **Table 1**.

**Table 1:** Demographic and clinical information of surveyed population (n=382).

| Characteristics | Value % (n) |
| --- | --- |
| Age (mean, SD) | 51.3 (11.4) |
| Gender |  |
| Male | 58.6 (224) |
| Female | 41.4 (158) |
| Higher education degree | 10.2 (39) |
| Master's degree | 5.2 (20) |
| Doctorate degree | 5.0 (19) |
| Medical specialty | 82.5 (315) |
| Pediatrics | 28.0 (107) |
| General medicine | 17.5 (67) |
| Traumatology | 13.1 (50) |
| Gastroenterology | 6.8 (26) |
| Allergy | 5.2 (20) |
| Gynecology | 3.7 (14) |
| Dermatology | 3.4 (13) |
| Critical care | 2.6 (10) |
| Other | 19.6 (75) |
| Work experience (years) |  |
| 1-5 years | 9.2 (35) |
| 5-10 years | 10.5 (40) |
| 11-15 years | 13.1 (50) |
| >20 years | 53.4 (204) |

### Knowledge about telemedicine

Around half of participants expressed to be lowly to very lowly familiarized with telemedicine technology (χ^2^(4) = 88.497, *p* = .000), its application in medicine (χ^2^(4) = 102.188, *p* = .000) and telemedicine tools (χ^2^(4) = 106.874, *p* = .000) (**Fig 1, S1 Table**). Roughly 3 out of 10 patients had an average familiarity with such topics while only around 20% where highly to very highly familiarized.

**Fig 1.**
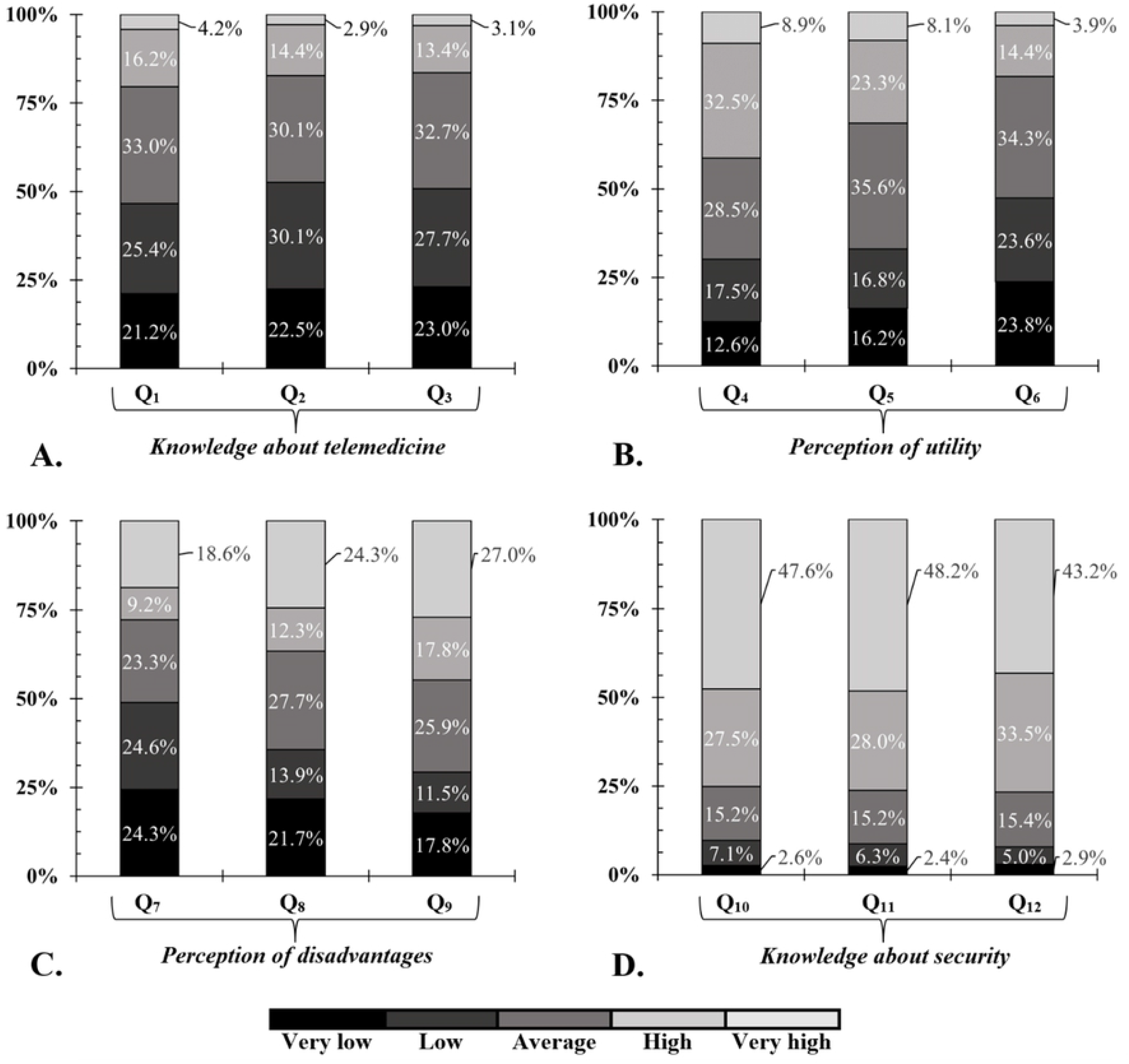
Proportions of categories expressing the extent to which participants asserted the influence of telemedicine on each of the questions encompassed by each domain. Q1, “ To what extent are you familiar with telemedicine technology?” ; Q2, “ To what extent are you familiar with the medical applications of telemedicine technology?” ; Q3, “ To what extent are you familiar with telemedicine tools?” ; Q4, “ In your opinion, to what extent is telemedicine effective in reducing the costs of patient care in hospitals?” ; Q5, “ In your opinion, to what extent does telemedicine technology save clinicians’ time?” ; Q6, “ In your opinion, to what extent does telemedicine technology provide faster and better medical care?” ; Q7, “ In your opinion, to what extent does telemedicine technology endanger patient privacy?” ; Q8, “ In your opinion, to what extent does telemedicine technology reduce the efficiency of patient care?” ; Q9, “ In your opinion, to what extent may telemedicine technology increase malpractice in healthcare?” ; Q10, “ To what extent should a framework be created to prevent breaching data confidentiality when using telemedicine?” ; Q11, “ To what extent does telemedicine technology require legal clarification for patients?” ; Q12, “ To what extent does telemedicine technology require a formulated and clear framework for access to medical information?”.

### Perception of the utility of telemedicine

Most of the participants (32.5%; n=124; χ^2^(4) = 78.812, *p* = .000) considered, to a high extent, that telemedicine is effective in reducing costs of patient care in hospitals. However, most participants also alleged that telemedicine technology can save an average amount of clinicians’ time (35.6%, n=136; χ^2^(4) = 80.277, *p* = .000) or provide, to an average extent, a faster and better medical care (34.3%, n=131; χ^2^(4) = 99.571, *p* = .000) (**Fig 1, S1 Table**).

### Perception of the disadvantages of telemedicine

Even though most participants reported that telemedicine technology may endanger patient privacy either to a very low (24.6%, n=94) or low (24.3%, n=93) extent (χ^2^(4) = 32.555, *p* = .000), most participants considered that it can highly increase malpractice in healthcare (27.0%, n=103; χ^2^(4) = 31.534, *p* = .000) (**Fig 1, S1 Table**). Also, 27.7% (n=106) expressed that telemedicine technology reduces the efficiency of patient care in an average extent while 24.3% (n=93) considered this statement to a high extent (χ^2^(4) = 31.534, *p* = .000).

### Knowledge of the security of telemedicine

About 8 out of 10 participants expressed that, from a high to a very high extent, a framework should be created to prevent breaches of data confidentiality when using telemedicine (χ^2^(4) = 250.749, *p* = .000), that telemedicine requires legal clarification for patients (χ^2^(4) = 263.628, *p* = .000) and that telemedicine technology requires a formulated and clear framework for access to medical information (χ^2^(4) = 240.670, *p* = .000) (**Fig 1, S1 Table**).

## Discussion

Telemedicine is a growing field with the potential to improve healthcare delivery, however it is important for a proper implementation to understand how physicians and patients perceive the benefits and limitations related to their use. In our study we found that roughly half of participants expressed a low familiarization with telemedicine technologies and its application in medicine. This finding contrast with a previous study among physicians in Saudi Arabia, in which around 46.1% reported average knowledge about telemedicine.(14) It is possible that organizational structure and culture affect health care providers’ perceptions of telemedicine based on the diffusion of innovation theory.(15) According to this theory, the key to adoption is that individuals must perceive the idea as “ innovative” to enable greater diffusion. This is one area where a change in healthcare organizations’ s culture and structure may be able to affect health care providers’ perceptions of telemedicine.(16)

Despite the low familiarization with telemedicine among physicians in our study, most of them considered to a high extent that it can be effective in reducing the costs related to patient care in hospitals, save a considerable amount of clinician’s time, and provide faster and better medical care. Similarly, a previous study among physicians in Indonesia found that most respondents considered that telemedicine is beneficial for patients (89%) and were interested in its continued use (88%).(17) Previous studies have shown that telemedicine can be particularly useful for patients with chronic diseases.(18) For instance, Nguyen and colleagues reported that after taking into consideration all costs and effects of a telemedicine-based national diabetic screening program in Singapore, it would still had significantly lower costs while generating similar quality-adjusted life-years compared to a physician-based model.(19) In a randomized control trial from Japan authors found that the use of a mobile application was a cost-effective tool that might help to reduce the incidence of dysmenorrhea and depression among women, and interestingly enough most participants were willing or relatively willing to use it.(20)

Despite its benefits, there are several aspects of telemedicine inherent to its nature such as security and privacy concerns, and the danger to affect patient privacy or incur in malpractice. In our study 4 out 10 respondents considered to a low extent that telemedicine may endanger patient privacy, however a similar proportion stated that it may increase malpractice cases. Additionally, 8 out of 10 participants in our survey expressed that a framework should be created to access medical information and prevent breaches of data confidentially when using telemedicine. These interesting findings may be the related to the fact that as of today, mobile technologies not only manage personal data but also highly personal information including social interactions and emotions, creating new issues such as the appropriateness of physicians to communicate with patients through platforms outside of the electronic medical record.(20) This represents an obstacle that needs to be addressed by healthcare organizations and providers to ensure that sensitive information is only shared among authorized individuals.(21,22)

Prior studies have determined that there is a growing interest in the use and application of telemedicine in the clinical practice, but there are still barriers ranging from the availability of internet and mobile devices to limitations of physical assessments, and even resistance to change that can affect the experience and satisfaction of using technology on a case-by-case basis.(23,24) As of today, telemedicine represents a primary means of expanding care to those with limited access to physicians, but to be truly patient centered it must also be affordable and accessible.(25) Considering the perceptions and knowledge of physicians related to telemedicine is also essential to design more efficient and easier to use tools that ultimately will increase the participation in these new technologies.(8)

## Limitations

In light of our findings there are several limitations worth mentioning. Nearly one third of our sample consisted of pediatricians, while a sixth of it was composed of traumatologists, which may limit the generalizability of our results to physicians with other specialties. Around half of the participants were unfamiliar with telemedicine, which may lead to biased answers when asked questions about topics requiring more in-depth knowledge of these technologies. However, to the best of our knowledge our study is among the first to assess the perceptions of telemedicine among Ecuadorian physicians, providing valuable insights that might be useful to design future interventions.

## Conclusion

Understanding physician’s perceptions to the use of technology is an important step for identifying unmet needs and areas of improvement. In this study we found a considerable proportion of physicians reporting low familiarization with telemedicine despite being aware of the benefits it can bring to patient care. Breaches of data confidentially and the potential for malpractice were cited as main concerns in need of a framework to prevent them. Future studies are needed to address the perceived barriers of technology to ensure a safe and efficient use of telemedicine in the healthcare setting.

## Data Availability

The following supporting information can be downloaded at: https://docs.google.com/document/d/1ctHGh4VMW_zmbnTjiNnPfBH9EKJPMgKS/edit?usp=drive_link&ouid=105604320198849901573&rtpof=true&sd=true S1 Table: Percentages and frequencies of categories that indicate the extent to which participants asserted the influence of telemedicine on each of the topics covered by each domain.

## Acknowledgments

The authors acknowledge the guidance and knowledge imparted by the MECOR Program, especially Sonia Buist and Ana Menezes, for this study. Special thanks to all members of Respiralab Research Group in particular Dr. Christian Kuon Yeng for their initial input regarding this project. We want to also express our gratitude to Dr. Krunal Pandav (Larkin Health System, South Miami, FL, USA) for his early contributions in manuscript preparation. Finally, we want to express our gratitude to Universidad Espiritu Santo for their continuous support in our research endeavors.

## Supporting information

**S1 Table. Percentages and frequencies of categories that indicate the extent to which participants asserted the influence of telemedicine on each of the topics covered by each domain**. All data are presented as percentages and frequencies. Differences in values between the five “ extent” groups are significant at .05 significance level.

